# MUSCLE: Muscle Understanding through Synthetic Computation and Lesion Evaluation A Semi-Synthetic Dataset for Hamstring Injury Prediction Using Electrical Impedance

**DOI:** 10.1101/2024.11.12.24317096

**Authors:** Lea Youssef Baby, Reem Shehayib, Noel Maalouf

## Abstract

Hamstring Injuries (HSIs) are common among athletes and necessitate extended rehabilitation before Return to Sport (RTS). Post-injury, athletes undergo physical examinations, which often fall short in assessing injury severity or guiding rehabilitation. Therefore, imaging techniques such as Magnetic Resonance Imaging (MRI) are used to evaluate the injury more comprehensively, aiding in the assessment of optimal rehabilitation and RTS timelines. Given the significant impact of HSIs on athletic careers, early prediction is essential. This article investigates the use of Electrical Impedance Tomography (EIT) for HSI prediction. EIT, a noninvasive method, involves injecting a current or voltage into the affected area to detect property changes, allowing for real-time monitoring and supporting its role in HSI prediction. A semi-synthetic dataset was created using MRI scans of patients with hamstring injuries. The dataset was developed by mapping the boundaries of the hamstring muscles (semimembranosus, semitendinosus, and biceps femoris) with Electrical Impedance Tomography and Diffuse Optical Tomography Reconstruction Software (EIDORS). EIDORS generated EIT voltage measurements by defining muscle boundaries and setting appropriate properties, forming the basis for the dataset. Machine Learning (ML) models were then employed to validate the dataset by distinguishing between injured and healthy hamstrings. The best-performing model, Random Forest (RF), achieved an accuracy of 98%, demonstrating the potential of EIT in predicting HSIs.

Figure 1:
Graphical Abstract

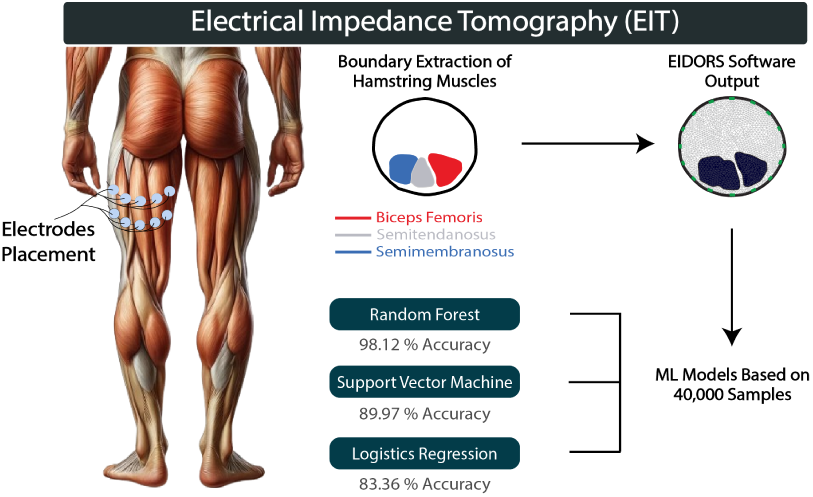

## 1 Introduction

HSIs are extremely frequent among athletes and require an extended duration of rehabilitation before Return to Sport (RTS). These injuries are accompanied by the intervention of rehabilitation practitioners to prepare athletes for RTS. After RTS, risks of re-injury are high, which compromises the performance of the athletes. Previous HSIs lead to a 2.4 times increase in chances of having another HSI in the weeks after RTS [1]. Ousmane Dembéĺe, a French soccer player born in 1997, endured a total of 7 HSIs dating back from 2017 until 2024 [2]. He faced multiple re-injury occurrences after extended periods of rehabilitation. An example of that is an HSI that he got on November 4th, 2021, after being in rehabilitation for 134 days due to a knee injury, which caused him a total of 18 extra days of rehabilitation while missing 4 games. Additionally, on February 4th, 2020, Ousmane Dembéĺe faced an HSI, which led to 191 days of rehabilitation and a total of 19 games missed [2]. The example of this athlete highlights the dangers and impact of an HSI on the time needed for rehabilitation before RTS and the enhanced risk of re-injury. Therefore, HSI prediction is extremely essential and needed to aid athletes in reducing their time of training due to rehabilitation and the multiple challenges that come along with an HSI, which is the contribution of this current study.

### 1.1 Anatomy of the Hamstring Muscles

The hamstring muscles are formed of three main muscles: the semimembranosus, the semitendanosus, and the biceps femoris. These muscles are located along the back of the thigh and originate from the ischial tuberosity. The biceps femoris has two heads, the long head and the short head. On one hand, the long head of the biceps femoris and the semitendanosus share a common tendon while the semimembranosus has a different thicker tendon. On the other hand, the short head of the biceps femoris is attached to the posterior femur. These muscles aim at flexing the knee and extending the hip. Each of these muscles also has unique attachments that permit complex movements. Additionally, the stability of the knee is due to the fact that the semitendanosus and the semimembranosus insert on the medial tibia while the biceps femoris inserts laterally on the fibula and tibia [3].

### 1.2 Causes and Treatment of Hamstring Injuries

HSIs are mainly caused by two different events. On one hand, they can be caused by repetitive damage to the muscle, which leads to an “ongoing decline of tissue integrity” [1]. When an HSI occurs, individuals feel a sudden “posterior thigh pain”, which is sometimes accompanied by an “audible or sensory pop”[1]. Sports that include fast acceleration, deceleration, and changes in direction, such as track-and-field, soccer, and American football, are common reasons behind a classical hamstring strain. Additional factors such as previous lower extremity trauma, older age, and deconditioning also contribute to the occurrence of HSIs. Typically, when one muscle is involved in the strain, it is mostly the long head of the biceps femoris, whereas the semimembranosus is considered the second most commonly affected. In one-third of the cases, more than one muscle is affected by the strain, and generally, it is the long head of the biceps femoris along with the semitendanosus or with the short head of the biceps. To treat muscle strains, typically, the “PRICE” principle is adopted. “PRICE” stands for Protection, Rest, Ice, Compression, and Elevation. Additionally, analgesics or nonsteroidal anti-inflammatory drugs are used for pain control. This treatment is then followed by mobilization, stretching, and flexibility training programs [3].

When individuals have HSIs, they undergo two different examinations, subjective and clinical. The subjective examination includes reviewing the patient’s history to check for past HSIs. It should also be noted if the patient had injuries at their lower back, hip or groin, and knee since it might alter the rehabilitation process. Clinical examination includes multiple tests which are palpation of the injured area, Range of Motion testing, and Strength Testing. However, for a better assessment of the injury, an MRI is required. MRI is used to evaluate the degree of tissue damage and the location [1]. On the other hand, HSIs are caused by a macro-traumatic event where the limits of the muscle-tendon unit are exceeded [1]. This is also known as a hamstring avulsion. Avulsions are considered less common than muscle strains. When an avulsion of the proximal hamstring tendon from the ischial tuberosity occurs, the conjoint tendon is usually affected more than the biceps femoris tendon alone, whereas avulsions with a semimembranosus origin are rare. This type of injury is mostly caused by water skiing. Additionally, a common injury occurs due to the violent flexion of the hip while the knees are locked in full extension. Proximal avulsions also take place when activities such as splits, ballet, and gymnastics happen. Typically, partial tears are less common than complete injuries with or without tendon retraction. In the cases where complete avulsions occur, surgeries are required, and the recovery time ranges from 3 to 18 months [3].

### 1.3 Imaging Diagnosis

Imaging techniques such as MRI scans and ultrasounds are extremely important for two main reasons. First, it is considered impossible to distinguish between a muscle strain and a muscle avulsion using physical examination alone. Second, imaging techniques are used to plan the treatment process. It is important since, through an MRI, a partial or a full tear can be identified, and whether or not the conjoint tendon, the semimembranosus tendon, or both are involved.

Cross-sectional imaging can then be used to identify the characteristics of a muscle strain. Usually, when muscle strains occur, a macroscopic hematoma will appear in or between the muscles as a concentrated mass. The echotexture of the hematoma varies depending on its age and the amount of internal liquefaction. The most frequent observations that can be extracted from MRI scans are high-signal-intensity edema and hemorrhages that are centered at the “main myotendinous junction”. Based on that, the severity of the injury is reflected on a scale of 1 to 3 on the MRI. Grade 1 is associated with a mild strain, all myotendinous fibers are intact but may appear distorted. Grade 2 represents a macroscopic partial fiber disruption and hematoma in the space left between the disrupted fibers. Grade 3 involves a full rupture of the myotendinous cross-section [3].

After highlighting the impact of hamstring strains on athletes, along with the extended rehabilitation period required, finding ways to prevent and predict these injuries becomes extremely important. To achieve this, real-time monitoring is crucial for alerting the athlete when an injury might occur, which can be accomplished using a technology called EIT.

### 1.4 Electrical Impedance Tomography

EIT, a method that revolves around injecting a current or voltage to the target area to determine its response and examine changes in its properties, is non-invasive [4–7]. EIT is also known as an “impedance-based imaging technique” [8] where in other words, it is used to determine the internal impedance at different locations [8]. This technique is widely used for multiple applications, including pulmonary imaging, brain imaging, breast cancer detection, monitoring neural and brain activities, and evaluating perfusion and cardiac function[9]. EIT measurements are usually taken by placing electrodes at the surface of the body, which can be either polarized or non-polarized. Polarized electrodes are stainless steel, conductive fabrics, and rubbers, while non-polarized electrodes are ECG-type silver-silver chloride (Ag/AgCl) electrodes [10]. Figure 2 is a representation of an EIT system that can be used for pulmonary imaging. The figure shows how the electrodes are placed around the torso to make the appropriate recordings.

**Figure 2:**
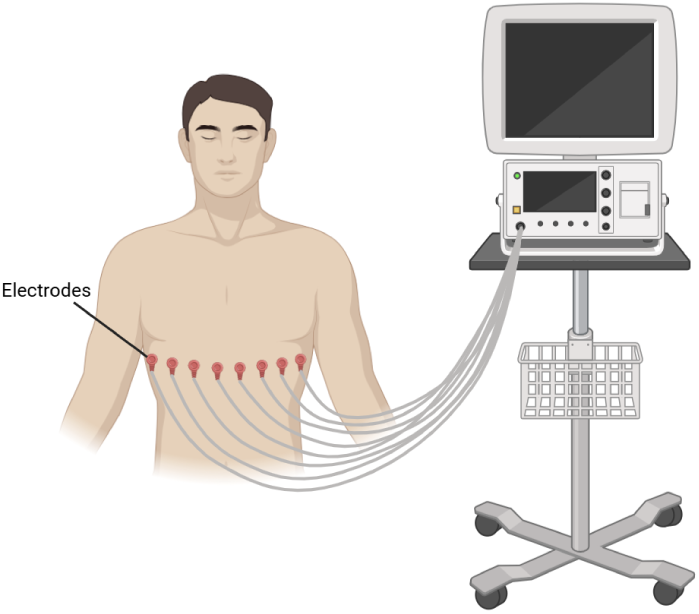
Representation of an EIT system used for pulmonary imaging, which consists of placing electrodes around the torso and recording EIT measurements. This image was produced using Biorender.

Ensuring the stability of the electrodes by integrating them into a belt or harness fixed around the target organs is essential since EIT is extremely sensitive, and having the smallest movement leads to major impacts on the quality of the signal[11]. The number of electrodes used dictates the precision of the constructed EIT image having a higher number of electrodes increases the precision of the image [11, 12]. At the electrode-tissue interface, gels, and liquids are also applied to ensure conductivity [11]. A decreased data quality is generally due to the dryness of the electrodes, motion artifacts, and changes in human posture. The injected waveform is usually sinusoidal or approximated by a square wave. It always follows the “electrical safety considerations” since it can vary in frequency, intensity, or nature (being a voltage or a current injection). The maximum allowed current to be injected into the body is 10 mA at 100 kHz frequency [13]. EIT is used to generate images that are 2D or 3D based on the sample at hand [8], but it presents two main problems. The forward problem and the inverse problem. On one hand, the forward problem consists of predicting voltage readings on a conductive object’s boundary when electrical currents are delivered to the electrodes. Then, the actual measurements obtained from the inverse problem when the internal conductivity is reconstructed are compared to these predicted voltages. Numerical techniques such as the finite element or the boundary element methods have been developed to estimate how the electrical potentials are distributed within the object [14]. On another hand, the inverse problem deals with reconstructing the internal conductivity distribution of an object from voltage measurements made at its boundary. This problem is ill-posed since small measurement errors lead to major reconstruction errors. Algorithms have been developed to solve this problem, such as the Gauss-Newton or modified Newton–Raphson. Then, an image of the internal conductivity fluctuations is obtained by solving it using the previously mentioned methods that iteratively modify an initial conductivity estimate in order to minimize the discrepancy between the observed and simulated (forward problem) boundary voltages [14] [15].

The obtained data needs to be processed and analyzed to calculate “application-relevant” images and measures since it is virtually impossible to give relevant clinical information based on raw data [16].

However, the use of EIT in muscle imaging is not yet widely explored. One of the main challenges and future goals in this research is to apply EIT for muscle imaging. Therefore this study makes the following contributions:

- Creation of a semi-synthetic dataset of EIT voltages for hamstring muscles based on MRI scans.
- Utilization of Electrical Impedance Tomography and Diffuse Optical Tomography Reconstruction Software (EIDORS) for the creation of the dataset.
- Validation of the dataset using ML models to show the effectiveness of utilizing EIT for HSI prediction.

The rest of the article is organized as follows. Section 1.5 highlights the state-of-the-art methods in predicting HSI and the current applications of EIT. Section 2 covers the methodology behind creating the dataset, followed by an overview of the extracted features from the EIT signals and the adopted machine learning models. Section 3 presents the results, which are discussed in Section 4, followed by potential future work and concluding thoughts.

### 1.5 State of the Art

Previous studies have been developed in an effort to predict injuries and optimize the rehabilitation process for athletes. A study conducted by Ĺopez-Valenciano et al. [17] explores multiple machine learning methods to identify the risk of having lower extremity muscle injuries (MUSinj) for athletes. Lower extremity muscles, including the adductors, hamstrings, quadriceps, and triceps surface, comprise more than 90% of injuries for soccer players [17]. This highlights the importance of having methods to predict MUSinj for athletes. To implement this strategy, a best-performing risk-injury factor model was developed. An experiment was conducted with a total of 132 professional male soccer and handball players. Soccer and handball are considered two very different sports but result in the high incidence rate of MUSinj. During the preseason phase, a set of tests was evaluated for all players, which included personal, psychological, and neuromuscular assessments, which are all considered sport-related injury risk factors. The preseason session tests were divided into three sessions with a total of 120 min. The first part is related to obtaining personal characteristics, sports-related background, and demographic features. The second part was focused on assessing psychological measures that correspond to sleep quality and athlete burnout. And, the third part was done to evaluate neuromuscular measures. It consisted of performing a set of exercises comprising dynamic postural control, isometric hip abduction and adduction strength, lower extremity joint ranges of motion (ROM), core stability, and isokinetic knee flexion and extension strength. A supervised learning perspective was utilized in order to compare the behaviors of multiple machine learning techniques, and the best model for the prediction of MUSinj was chosen. The process is divided into two parts: data pre-processing and data processing. Data pre-processing consisted of data cleaning and data discretization. While for data processing, the paradigms that were used are the ones that allow to deal with high-dimensional and imbalanced datasets. Several pre-processing methods and ensemble techniques were developed to classify players based on high to low chances of MUSinj. The best model used to predict athletes at high risk of MUSinj resulted in a 0.747 AUC (area under the ROC curve) score, 65.9% TPrate and 79.1% TNrate [17].

A similar study was conducted by Ayala et al. [18] to compare numerous Machine Learning (ML) models and select the best-performing injury risk factor model to predict athletes that are at high risk of HSI. The experiment was conducted based on a total of 96 male professional soccer players. The players underwent a set of pre-season evaluations along with the collection of personal, psychological, and neuromuscular measurements. The first section of the testing phase was focused on gathering the player’s personal or individual characteristics. A set of questions related to the athlete’s background, along with his demographics, were recorded. The second section assessed the psychological state of the player to determine the sleep quality along with the burnout of the athlete. And, the third section was focused on evaluating numerous neuromuscular measurements. Before they conducted the third section of the testing phase, a 15-20 minute dynamic warm-up was led by athletes whereas the neuromuscular assessment was executed 3-5 min after the warm-up. Additionally, all HSI occurrences were recorded and sent to the study group on a monthly basis. Predictive models were subsequently developed using supervised learning, with the recorded risk factors as input features and a discrete variable representing the presence or absence of HSIs as the target outcome. The development of the model followed two stages. The first stage involved data preprocessing, which included cleaning and discretizing the data. The second stage focused on data processing, using Decision Tree (DT) algorithms as the basis for the various models tested. The best-performing model achieved an AUC of 0.837, a true positive rate (TPrate, also referred to as sensitivity or recall) of 77.8%, and a true negative rate (TNrate, where actual negatives are correctly classified as negative) of 83.8%. Despite the model’s success in making accurate predictions, its high complexity limits its ability to explain why an HSI occurred [18].

In the process of predicting the injury recovery time, Skoki et al. [19] develop ML models which depend on the physician’s perspective. The models developed were the following: linear regression, DT, and extreme gradient boosting (XGB). These models were trained using a total of 69 injuries of male professional soccer players. The features used in the model included information about the injury, whether it resulted from a tackle, and the injury classification based on a scale from 0 (minor injury) to 4 (major muscle rupture) according to the BAMIC (British Athletics Muscle Injury Classification) grading system, as well as the position, depth, and location of the injury. Additional features incorporated general, injury-specific, and recovery-related information. It was shown that adding the expert’s prediction as a feature improved the model’s performance, resulting in a best-performing XGB model with a mean R^2^ of 0.72953. This performance can be compared to the expert’s own prediction, which achieved a mean R^2^ of 0.62242 [19].

Caldeón-Díaz et al. [20] focus their study on developing an ML model used to determine professional soccer players at high risk of having HSIs. This is done by using multiple ML models, including DT, Logistic Regression, discriminant method, Support Vector Machine (SVM), and others. Additionally, XGBoost is used to determine the most important features contributing to a correct prediction. Through the feature importance module of the XGBoost, it was determined that the features contributing the most are the stiffness and the maximum muscle strength of the hamstring muscles. After testing 35 different ML models, the best-performing model achieved a 78% precision showing the reliability of the model to assist doctors in predicting HSIs. The dataset used for training the ML models is based on 110 male professional players where the data collected from each player is the age, weight, height, biomechanical test results, anthropometric measurements, and the position of the player in the team. This data results in a total of 19 different features used to train the model. Pre-processing was then applied to the data followed by a classification process having a 110 x 19 feature matrix. The models were categorized into two classes: class 0, indicating no lower limb muscle injury, and class 1, indicating the presence of a lower limb muscle injury. [20].

Since the choice of the injury risk factors is not agreed upon, Henriquez et al. [21] developed an RF model to optimize the choice of these factors. Another model is then created for lower extremity musculoskeletal injury risk for student-athletes since it is reported that 90% of student-athletes face athletic injuries. The study conducted constituted 122 college athletes aged on average 19.56 *±* 1.33 years. The data used to train the model is based on 50 physical metrics of strength, postural stability, and flexibility assessments, along with demographic data and a binary classification of previous injuries. The strength assessment is determined using force transducers which evaluate joint strength. Postural stability assessments are tested using a force plate for both static and dynamic states. Finally, the flexibility assessment is obtained by examining the joint range of motion and muscle-tendon by using a digital inclinometer or a standard goniometer. Afterwards, RF algorithms were used to determine the most important injury risk factors which were picked based on the mean decrease accuracy. The mean decrease in accuracy highlights the decrease in accuracy if a variable is removed from a feature when the model is developed. Additionally, a model of lower extremity musculoskeletal injury risk is implemented while having the variables that resulted in a negative mean decrease accuracy removed from the final model. The output of the model is represented by the probability of the individual being injured. However, it is almost impossible to predict an injury based on risk factors since a student might have a high-risk factor but conducts successful training, which reduces the chances of being injured in comparison to another student who can have a very low injury risk but can get injured by undergoing an aggressive contact injury [21].

Therefore, another way to assess muscle health and thus be able to predict muscle injuries is through bioimpedance (BI) measurements. A study conducted by Nescolorade et al. [22] highlights the importance of using BI measurements in classifying skeletal muscles as healthy or injured. Specifically, the localized-bioimpedance (L-BIA) method is used to assess muscle injury and differentiate between tissue damage and localized fluid accumulation. Additionally, the importance of L-BIA can be seen in determining the (Return To Play) RTP status for athletes. The paper discusses how to determine the Resistance (R), the Reactance (Xc), and the phase angle (PhA), which are used to assess the physiological state of the muscles. To determine these parameters, it is noted that the human body can be mapped as a combination of resistors and capacitors and, thus, can be represented as a resistor-capacitor (RC) circuit. R is associated with the conductive properties (water and electrolyte), and Xc is related to the capacitive properties (cell membrane and tissue interface). An Alternating Current (AC) is introduced to the body, which splits it into two different paths: conductive and capacitive. Since the current is alternating, R is defined due to any impairments in the current flow. Additionally, the presence of non-conductive elements (cell membranes) leads to a phase shift between the current and the voltage due to the time delay that occurs which is due to the capacitive property of the tissues. To reach good results using L-BIA, the correct placement of the electrodes is essential. This placement differs based on the injury configuration. A short configuration (where the injury is far from the bone) dictates placing the voltage electrodes 5 cm proximal and 5 cm distally. A long configuration (where the injury is close to the bone) dictates placing the voltage electrodes 10 cm proximal and 10 cm distally. In either of these configurations, the current injection electrodes are placed close to the voltage electrodes. L-BIA can also overcome a limitation faced by radio-logical imaging criteria from MRI and US scans, which is their ability to predict the rehabilitation period for athletes. In sports-related injuries, the RTP is dictated by the severity of the injury itself which can be associated with the muscle gap interpreted as the retraction of muscle fibers. The retraction can be seen mostly in MRI scans. Additionally, the recovery time is also associated with the amount of edema in the injured muscle. Through L-BIA, the values of R, Xc, and PhA can be used to determine the severity of the injury, and it was proven that these values are inversely related to the injury severity [22]. A study was conducted with three professional soccer players where the BIA measurements were determined for quadriceps, hamstrings, and calf muscles before and after injury until RTP. It was proven that R, Xc, and PhA decrease with the severity of the injury. Additionally, the decrease of R is associated with fluid accumulation, while the decrease of Xc and PhA is linked to a disruption of cellular membrane integrity and injury [23]. A similar study conducted by Nescolarde et al. [24] also proves the correlation between the BIA parameters and the severity of the injury. BIA was performed at 50kHz 24h post-injury to assess both the integrity of the muscle structure and the fluid accumulation for 21 injuries at the levels of the quadriceps, hamstrings, and calf, which were also diagnosed using MRI scans. It was shown that the decrease in the R parameter was proportional to the presence of fluid distribution in the muscle but was not related to the severity of the injury, unlike the Xc parameter, which was shown to decrease with the increased severity of the injury [24]. These studies prove that using bioimpedance signals is an effective method to assess muscle injuries. The combination of bioimpedance signals, namely Electrical Impedance Myography (EIM) signals with ML techniques, can be used for clinical predictions. EIM is a non-invasive method that consists of placing typically 4 electrodes to minimize polarization and maintain a good signal quality on the tackled area. The outer electrodes apply an alternating sine wave current in the range of kHz to MHz that stimulates the tackled tissues. The tissue then generates an alternating voltage signal which is sensed by the inner electrodes. Figure 3 is a representation of the described EIM model.

**Figure 3:**
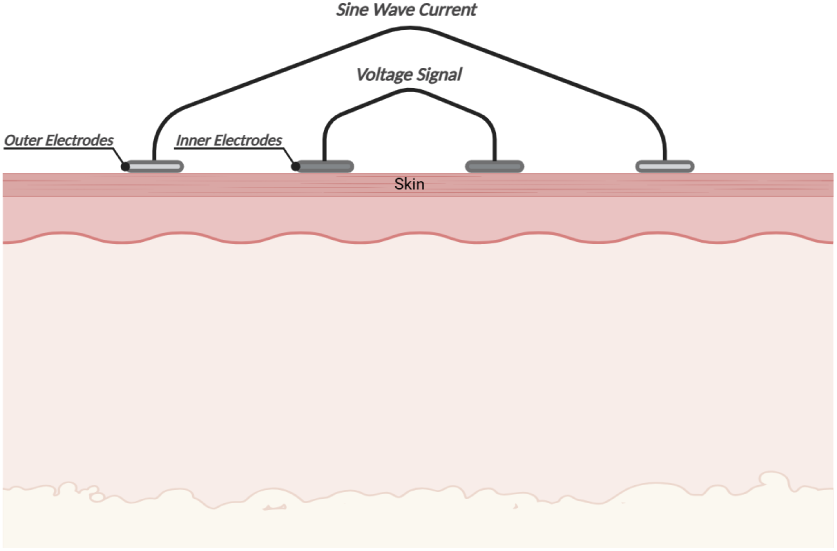
Representation of an EIM model. 4 electrodes are placed at the surface of the skin, having the outer electrodes apply a sine wave current signal and the inner electrodes sensing the generated voltage signal. This image was produced using Biorender.

EIM can be used to assess neuromuscular diseases, their severity, progression, and their response to therapy [25]. Additionally, EIM can also be used to evaluate changes in the muscle architecture. This includes myocyte atrophy and loss, edema, reinnervation, and deposition of endomysia connective tissue and fat [4]. Cheng et al. [26] discussed using ML models to estimate the total mass of thigh muscles (MoTM) based on EIM measurements and general body information. A total of 96 subjects participated in this study, 42 males and 54 females. The body information used to train the ML models includes the age, height, weight, BMI, and the circumferences of the thigh and calf. The performance of the models was evaluated based on the regression coefficient (r^2^) and the Root-Means-Square-Error (RMSE). The parameter r^2^ had a value of 0.800, and RMSE had a 0.929. This proves the effectiveness of using ML models with EIM measurements to predict the MoTM [26]. A different approach to assessing muscle health is through Electromyography (EMG). Specifically, a study conducted by Massó et al. [27] discusses using Surface Electromyography (SEMG) in sports. SEMG allows the determination of the electrical signals from a muscle in motion and has many applications. These applications include the analysis of movement and gait, the evaluation of fatigue and muscle activity through a diagnostic or therapeutic session, the assessment of neuromuscular disorders, and the evaluation of sports performance. The use of SEMG in sports is effective due to the feature of using SEMG in a dynamic state. Additionally, this technology is in practice in the sports field since it helps in the amelioration of physical activity by analyzing the activation of the muscles and determining muscle fatigue. Several steps are followed to determine EMG signals. The first step involves preparing the individuals, which includes informing the participants about the procedures to be followed during the experiment and obtaining their written consent. This step is followed by the preparation of the skin where an abrasive gel is applied to reduce the dry layer on the skin and thus reducing the impedance of the skin. Afterward, the positioning of the electrodes is extremely important to have good results. The electrodes should be placed on the midline of the belly of the muscle. Then, the Maximum Voluntary Contraction (MVC) is obtained to be able to compare the recordings between different subjects. Recording of the SEMG signal is then determined after having the participants execute a set of actions. Finally, signal processing is applied to enable proper analysis. The processing includes filtering the signal, rectifying it, applying a smoothing algorithm, and standardizing it according to the MVC [27]. Farina [28] conducted a study focused on techniques used to analyze EMG signals in dynamic contractions. Three main factors make the EMG signals in a dynamic setting challenging to analyze. These factors are signal nonstationarity, electrode shift, and the conductivity of the tissues. Signal nonstationarity occurs since, in a dynamic setting, there is a fast rate of change in the signal properties due to the changes in joint angles and the recruitment/de-recruitment of motor units. The problem related to the electrode shift is that measurements differ based on the positioning of the electrodes. Thus, by having a change in the joint angles, this positioning might change, which causes a change in the signal. Finally, tissue conductivity changes, as it depends on the muscle fiber orientation, as well as their lengths and diameters. Based on the type of information that needs to be extracted from an EMG signal the techniques to analyze dynamic SEMG differ. There are two main categories of required information: the degree of muscle activation and the properties of the membrane muscle fiber. Two main conditions need to be satisfied to successfully detect the activation of the muscles: (1) the absence of an EMG signal when the muscle is not active and (2) the ability to distinguish between noise and the EMG signal when the muscle is active [28].

Several methods have been developed to classify EMG signals. A study conducted by Yousefi et al. [29] has been developed for that purpose. An Artificial Neural Network (ANN) is a tool for classifying EMG signals. The reason behind the use of ANN is that they can be trained on a large dataset having high features and their learning capabilities which is needed to analyze and understand EMG signals. The only limitation mentioned is that it is not easily explained how the ANN models achieve their results [29]. Pattichis et al. [30] focused their work on the development of a model used to diagnose neuromuscular diseases using EMG. This was achieved by using ANN along with a Pattern Recognition Model (PRM). Supervised and unsupervised training paradigms were used. The model was based on data acquired from 44 individuals, 14 healthy patients, 14 myopathic patients, and 16 neuropathic patients. The signals were from the biceps brachii muscles after having the individuals perform voluntary contractions. The best-performing model reached a high accuracy of 80-90% [30]. As seen, multiple datasets have been developed in an effort to predict muscle injuries using different approaches, such as determined body information using BI, EIM, and EMG. However, no dataset is available for EIT measurements that can be used to predict muscle injuries. Examples of readily available EIT datasets include EIT measurements for stroke patients or measurements determined from a tank filled with saline solution.

A study conducted by Goren et al. [31] focused on using Multi-Frequency EIT (MFEIT) collected from 23 stroke patients and 10 healthy patients to create a dataset aimed at classifying strokes. 31 current injections were performed at 17 different frequencies ranging from 5 Hz to 2 kHz where each frequency yielded 930 measurements. This dataset allows the identification between healthy and stroke tissues. The results of the study show MFEIT’s potential in non-invasive stroke classification and its contributions to the creation of diagnostic instruments for early neurological injury diagnosis and real-time monitoring [31]. Additionally, Bounik et al. [8] discuss the importance of using multi-frequency impedance tomography to examine frequency-dependent characteristics. Due to the high complexity of such measurements, using ML models is an efficient approach to analyze these measurements, especially when EIT is used for imaging purposes [8].

Another study conducted by Hauptmann et al. [32] focused on producing an EIT dataset aimed at aiding researchers in improving EIT image reconstruction. Data was collected from a tank filled with saline water that is equipped with 16 electrodes. Various conductive and resistive objects were used to simulate different real-world imaging settings. Various EIT methods can be used to test this dataset and improve image quality which highlights its usefulness [32].

## 2 Materials and Methods

### 2.1 Creation of the Dataset

The semi-synthetic dataset is created using EIDORS [33]. EIDORS is an open-source software tool developed on MATLAB that is used for image reconstruction based on EIT measurements. It is used in multiple fields, including biomedical, industrial, and geophysical. The creation of the EIT dataset for HSIs is based on an innovative approach developed by Seo [34] that consists of feeding the boundaries of the targeted organ (namely the lungs) along with its electrical properties to the software. The software then generates a meshed output and a set of voltages representing the EIT measurements [34]. Therefore, the basis of this research is to adapt the work done for the lungs to the hamstring muscles. This can be done since, after a muscle injury, muscle fibers retract, which is seen as a muscle gap, and thus, a modification of the muscle area occurs [22].

### 2.2 Boundary Extraction

The first step to develop the dataset is to determine the boundary points of the hamstring muscles-which, as previously mentioned, are formed of three muscles, the semimembranosus, the semitendanosus, and the biceps femoris - as well as of the thigh seen from an axial view. The best way to get a detailed axial view of the hamstrings is to use MRI scans. However, finding publicly available datasets of injured hamstring muscles is challenging. Therefore, a total of 6 MRI scans were used to generate the semi-synthetic dataset. Table 1 highlights the different MRI scans used. The scans range from grade 1 to grade 3 injuries, where the chosen MRIs have injuries that impact the 3 muscles of the hamstrings. Specifically, 3 scans were taken from the study conducted by Rubin [3], where MRI 01 from Table 1 is for a male patient in the age range of 21-25 years with a grade 1 distal myotendinous hamstring strain. The edema is centered between the distal muscle belly and the tendon of the semitendanosus. MRI 02 also highlights a grade 1 injury where the edema covers 10% of the lateral margin of the biceps femoris. The scan is of a male patient in the age range of 26-30 years. MRI 03 from Table 1 is of a male athlete patient in the 31-35 years age range. The athlete got his injury from pre-race stretching. The scan shows edema that is throughout the semimembranosus muscle with surrounding fascial edema [3]. MRI 04 shows the injury of a male patient who belongs to the 51-55 age group with a grade 3 injury due to a torn semimembranosus. The patient got injured after falling which resulted in pain in the back of his thigh. The injury led to a high-grade muscle tear of the semimembranosus [35]. MRI 05 is from a male patient who belongs to the 61-65 age group and got injured while exercising, which also resulted in pain in the back of the thigh in the region of the hamstring muscles. This injury is described through a tear of both the semimembranosus and the biceps femoris. Finally, MRI 06 is of a grade 2 injury at the right proximal semimembranosus strain [24].

**Table 1:**
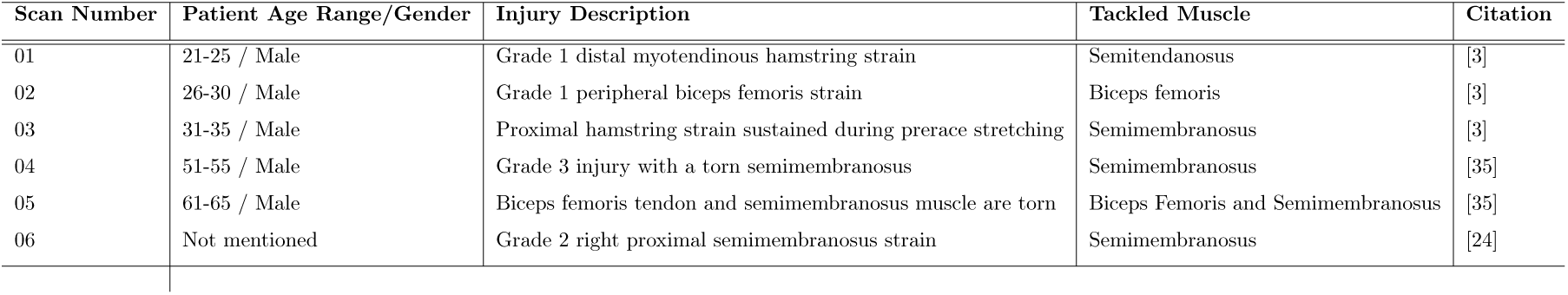
Summary of MRI Scans.

For each MRI scan, 20 different cases were generated, 10 of which represent healthy hamstrings and the other 10 represent injured hamstrings. In MRI scans, an injury can usually be detected by the presence of edema - around or across the muscle - or the retraction of muscle fibers. The intensity of the edema and the percentage of the cross-sectional area it covers can be used to assess the injury [3]. On one hand, to generate a healthy case of the hamstrings, the whole area of the muscles is considered as if no edema or muscle retraction is present. On the other hand, to simulate injured scenarios, the boundaries determined consider the whole area of the muscle tissues alone without taking into consideration the swelling and fluid buildup represented by edema. In his study, Rubin [3] showed that the edema covers between 10% and 50% of the muscle area [3]. Thus, to generate the different cases of injury for each MRI scan, the muscle areas are modified so that the edema covers 10% to 50% of the muscles. Consequently, this modification of area is the reason behind creating a semi-synthetic dataset.

The next step after obtaining the MRI scans is to determine the boundary points of the hamstring muscles and thigh contour. Algorithm 1 highlights the process of saving the boundary points. The image is first loaded, and the right width and height are set. Then, the MRI is displayed with the origins of the set axis being at the bottom left of the image. Capturing the boundary points from the user is enabled, which allows the user to select the points that need to be saved. Finally, these points are saved to a MATLAB file. Additionally, specific guidelines were followed to extract the boundary points. A total of 20 to 25 points should be selected to save the boundaries of the muscles, while 40 to 45 points should be used to save the boundaries of the thigh contour [34].

**Algorithm 1.**
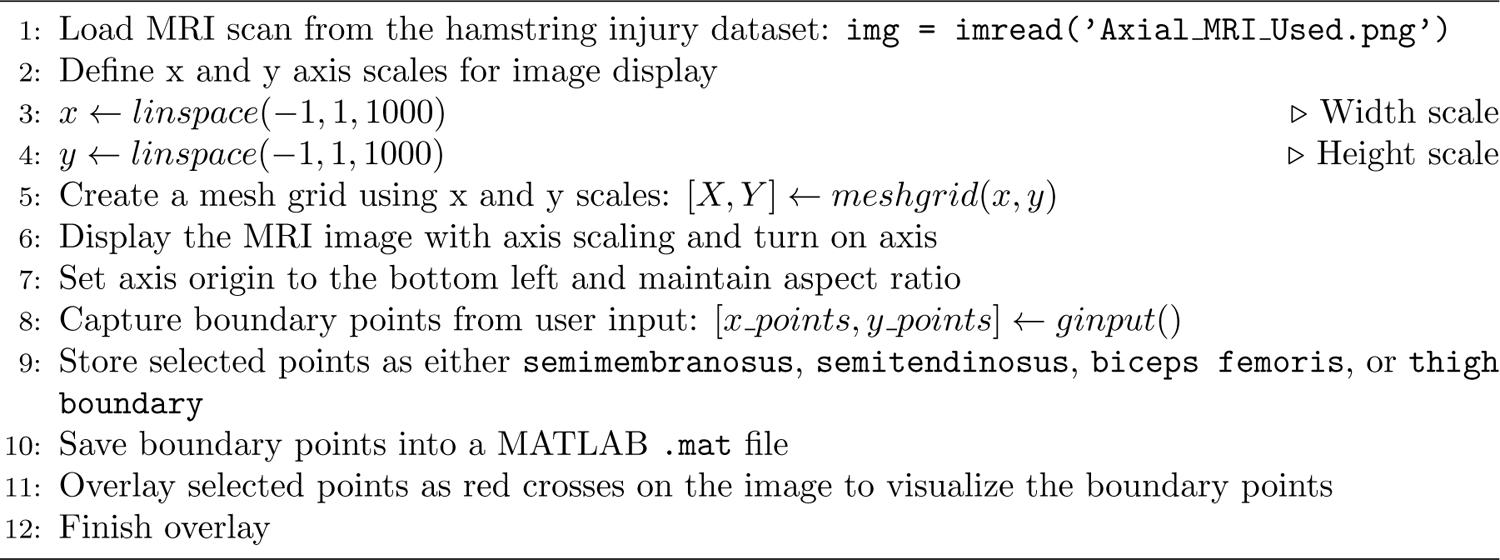
Determining the Boundary Points of the hamstring muscles and thigh After getting the necessary points for the muscles, it is important to save them in the *“shape library.mat”* that is provided by EIDORS. To do so, the data should be loaded into a variable, appended to the *“shape library.mat”* and then saved.

### 2.3 Generating EIT measurements using EIDORS

The next step after saving the boundary points of the muscles and the thigh contour is to feed them as inputs to the EIT EIDORS model created. Algorithm 2 shows the logic followed to determine the EIT voltages for an injured or healthy case. Initially, an EIT Data folder is created to store the files containing the voltage measurements. Next, the contours of the hamstring muscles and of the thigh are loaded. The shape of the model is then defined, which highlights the overall structure of the used FEM (Finite Element Model). The model is a 2D model with set smoothing parameters depending on the complexity of the shapes. The electrode shapes and positions are also defined. A total of 16 circular electrodes are placed at the boundary of the thigh on a single z-plane and with an equal distance between them. Then the FEM model is created and centered using netgen - an advanced mesh generator - by passing the defined parameters. The model is displayed and can be seen in Figure 4 for a healthy case of MRI 06 from Table 1.

**Figure 4:**
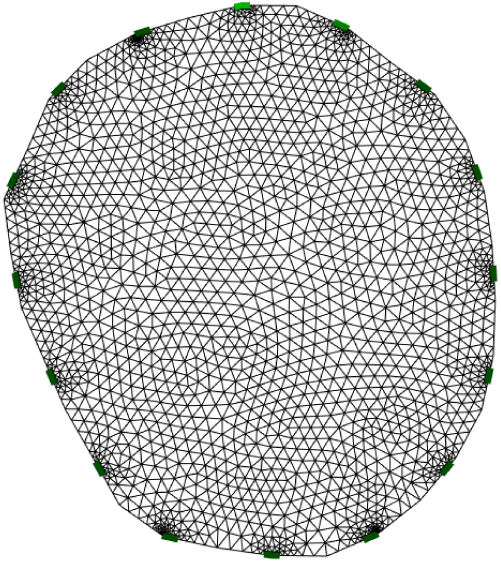
Creation of the FEM model using EIDORS after placing 16 electrodes at the boundaries.

The simulation patterns are then defined, showing how the currents and voltages are measured and applied across the electrodes. This is an essential step in EIT to solve the forward problem and determine the reference voltages. Then, the conductivity of the hamstring muscles is set depending on whether the simulation is for a healthy or injured case. A study conducted by Lee et al. [36] focused on determining in vivo measurements of thigh muscle conductivity. The study showed that the conductivity of healthy thigh muscles ranges from 1.15 to 2.58 S/m [36]. To simulate an injured hamstring case, a slightly higher conductivity range is considered since when an HSI occurs, a concentration of fluid in the muscle area happens which increases the ability of the muscle to conduct electricity. Afterward, a sine wave is generated to simulate the changes in the conductivity of the hamstrings.

The GREIT (Generalized Reconstruction for Electrical Impedance Tomography) model is defined to solve the inverse problem of EIT and reconstruct the image of the internal conductivity based on the results from the forward problem, which are the measured voltages. To accurately represent the shape of the model, it is essential to identify the boundary elements of the FEM model. Additionally, these boundary elements are used to convert from a triangular mesh to a grid mesh. This is done to ensure that the model is properly visualized and interpolated. To simulate different conductivity patterns, which can be due to a contraction of the muscles, an iteration over the defined sine wave is done. In that case, the FEM model is updated by giving it the new conductivity value, and the forward model is solved again. Solving the forward problem results in obtaining the new voltage data at the electrodes. GREIT is then applied to get a reconstructed conductivity image using the new voltages calculated. This step is followed by a conversion to a grid mesh. Post-processing is then applied to guarantee that the relevant internal region is considered while masking the outer body areas. Finally, the voltage data is saved to a CSV file which is then used to analyze the EIT measurements conducted. Figure 5 shows the final reconstructed image from the EIDORS software. For each of the 6 MRI scans, 20 distinct cases are generated: 10 representing healthy hamstrings and 10 representing injured hamstrings. Each case yields 1,000 CSV files containing voltage data, resulting in a comprehensive dataset of 120,000 voltage files. This dataset effectively simulates diverse EIT measurements across both healthy and injured hamstring conditions.

### 2.4 Feature Extraction

After determining the EIT voltage measurements, it is essential to extract features from the signal to improve the performance of the ML models developed. Testing on raw data with a low variety of essential features has resulted in poor performance and overfitting among the ML models. Thus, it would indubitably be game-changing to extract the appropriate features.

A total of 16 features were extracted and are the following: linear envelope, Root Mean Square (RMS), integration, zero crossings, Fourier coefficients, max and min values, variance, waveform length, autoregressive coefficients, temporal moment, Average Amplitude Change (ACC), V-order, log detector, Modified Mean Absolute Value (MMAV), and skewness. The extracted features are both in the time and frequency domains. The time domain analysis shows the variance of the signal with time, whereas the frequency domain analysis highlights the rate of change of the signal values [37]. Understanding each feature is essential.

1. Linear Envelope: It gives as a final result the outer shape of the signal after rectifying it and filtering it. The rectification is done by applying a full-wave rectifier, while the filtering is done by applying a Low-Pass Filter (LPF). It allows the signal to be smoother and thus facilitates its processing [38]. Figure 6 shows a plot of the linear envelope feature. The values of which were normalized to avoid having a bias while training the ML models.
2. RMS: It consists of finding the average power of the signal. To do so, first, the voltage values are squared which corresponds to the modification of power with time, then the obtained power waveform is averaged and the square root of the averaged power is obtained [39]. Figure 7 shows the plot from the RMS feature.
3. Integration: It determines the area under the signal curve after applying a full-wave rectifier [38].
4. Zero Crossings: It counts the occurrences of the signal crossing the zero line over a predefined window size [38].
5. Fourier coefficients: Since the analysis of signals in the frequency domain is an important aspect of processing and understanding the signal, Fast Fourier Transform (FFT) was applied to determine the Fourier coefficients [40] [38]. This feature highlights information related to the frequencies that are present in the signal along with their ratios [37].
6. Max and Min values: The maximum and minimum values from the signal are determined using these two features after applying an LPF.
7. Variance: It measures the degree of fluctuation of the signal in comparison to its mean value.
8. Waveform Length: It calculates the cumulative length of the raw, unfiltered signal [41].
9. Auto Regressive Coefficients: It consists of predicting the future values of the signal based on the current or previous values. Additionally, this feature allows for the reduction of signal noise and the enhancement of the signal’s resolution, spectral peaks, and data compaction [37].
10. Temporal Moment: It is a statistical analysis method that can be utilized as a feature [41].
11. AAC: It is close to the waveform length feature, but the difference is that the average of the wavelength is determined [41].
12. V-Order: The muscle’s contraction force is estimated using this feature which is a non-linear detector [41].
13. Log Detector: Similar to the V-order, the log detector also estimates the muscle’s contraction however, the non-linear detector is described in function of the logarithmic detector [41].
14. MMAV: It is considered as an extension of the Mean Absolute Value (MAV) of the signal; however, the weighted window function is added to the equation, which allows the assignment of different weights to the different portions of the signal. This approach offers more robustness to the MAV [41].
15. Skewness: It measures the asymmetry of a certain distribution where in that case, it evaluates the asymmetry of the EIT signal [42].

**Figure 5:**
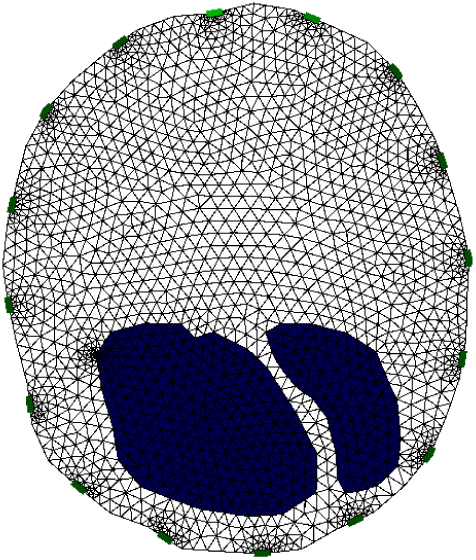
EIDORS model image of MRI 06 for a healthy case. The image illustrates the reconstructed image based on the EIT measurements generated by EIDORS.

**Algorithm 2.**
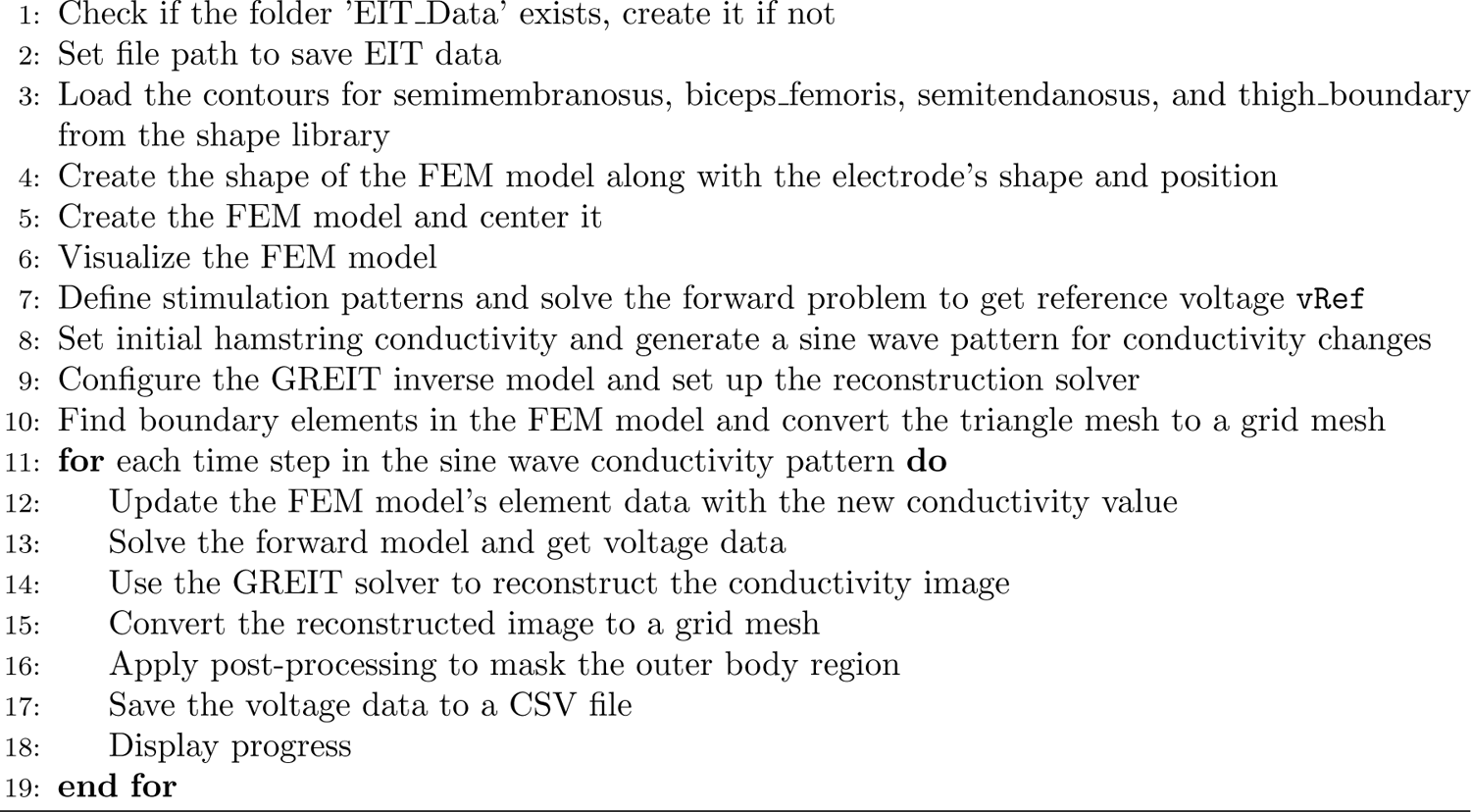
Obtaining EIT Voltages.

**Figure 6:**
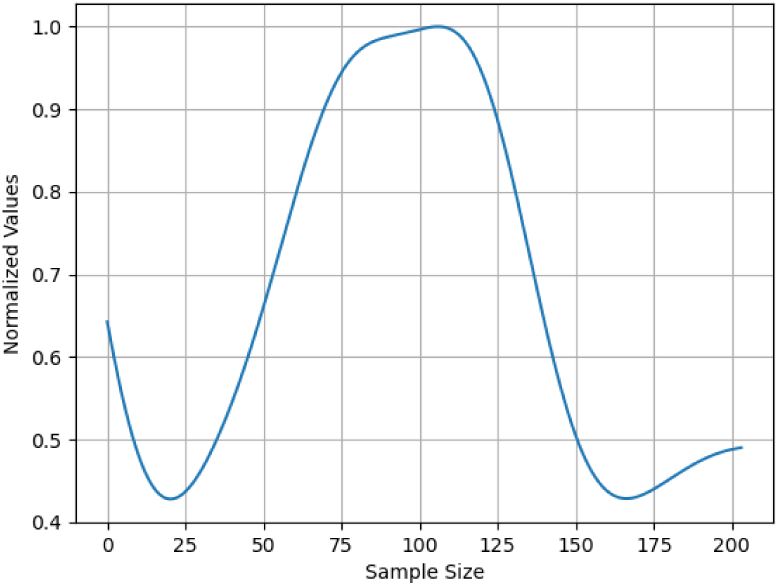
Linear Envelope feature extracted from the signal. The values on the x-axis represent the sample number, and the ones on the y-axis represent the normalized values of that feature.

### 2.5 Testing the Dataset

Utilizing machine learning (ML) models serves not only to achieve the objective of injury prediction but also to validate the dataset’s credibility. Accordingly, the complete dataset, comprising 40,000 samples, will be tested on three different generic ML models. Despite the sufficient sample size, we ensured that the data used to build these models accurately represented the original distribution. This alignment is crucial, as a classifier’s performance depends more on how well the dataset reflects the original distribution than on the sheer number of samples available [43].

### 2.6 Features’ Study

Other than having an appropriate collection of features that would positively affect the performance of our ML models, it is crucial to scrutinize the prominent features that solidify our data. To quantify the muscle activity through the signal over time, features such as *Root Mean Square (RMS)* and *Linear Envelope* are known to be frequently used [44]. After processing the raw signal, other features also include the *number of zero crossings* [44]. Furthermore, the *mean absolute value (MAV)* and *enhanced mean absolute value (EMAV)* constituted with the aforementioned features is considered to result in better performance, especially if combined with enhancement approaches [45]. Figure 8 displays the histograms of these features in addition to the *status* feature, which shows the equal distribution of both (healthy and injured) cases. The *rms feat*, representing the RMS features, appears to be approximately normally distributed with a peak around a central value, suggesting consistent measurement across the dataset. The *zero crossings* feature shows a slightly left-skewed distribution, with most values clustering at the lower end of the range, implying that the signal has fewer frequency changes or less noise. The *linear envelope*, labeled as *linear env*, can be considered to have a right-skewed distribution. This shows that there are significant occurrences of higher values while most data points have a low linear envelope value, indicating that there is increased signal intensity at some points. Regarding the *MAV*, we have it as *modified mav* feature, and it is considered somewhat normally distributed but with slight skewness to the right, thus indicating occasional spikes in amplitude. The *fourier series* feature is clearly right-skewed. This data distribution could be further useful in the future if identifying dominant frequencies or occasional high-energy bursts in the signal would be significant.

**Figure 7:**
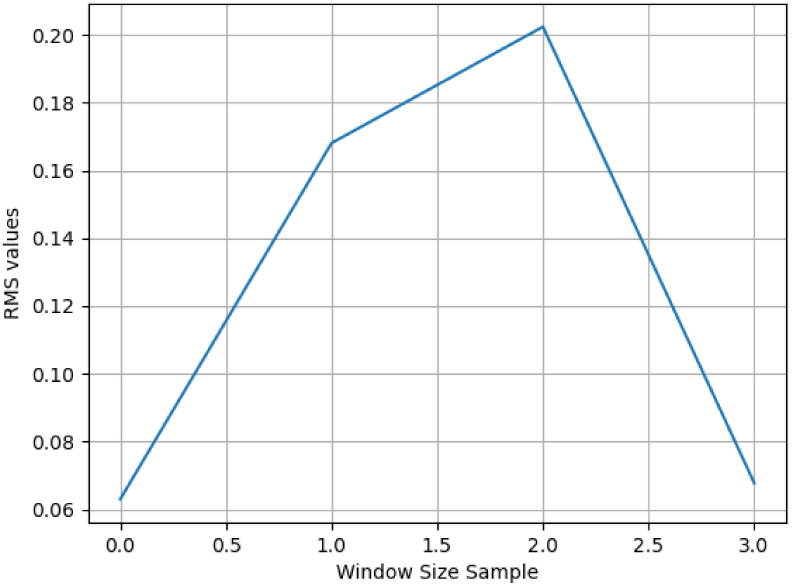
RMS feature extracted from the EIT signal. The plot represents the RMS values of the signal across a predefined window size.

**Figure 8:**
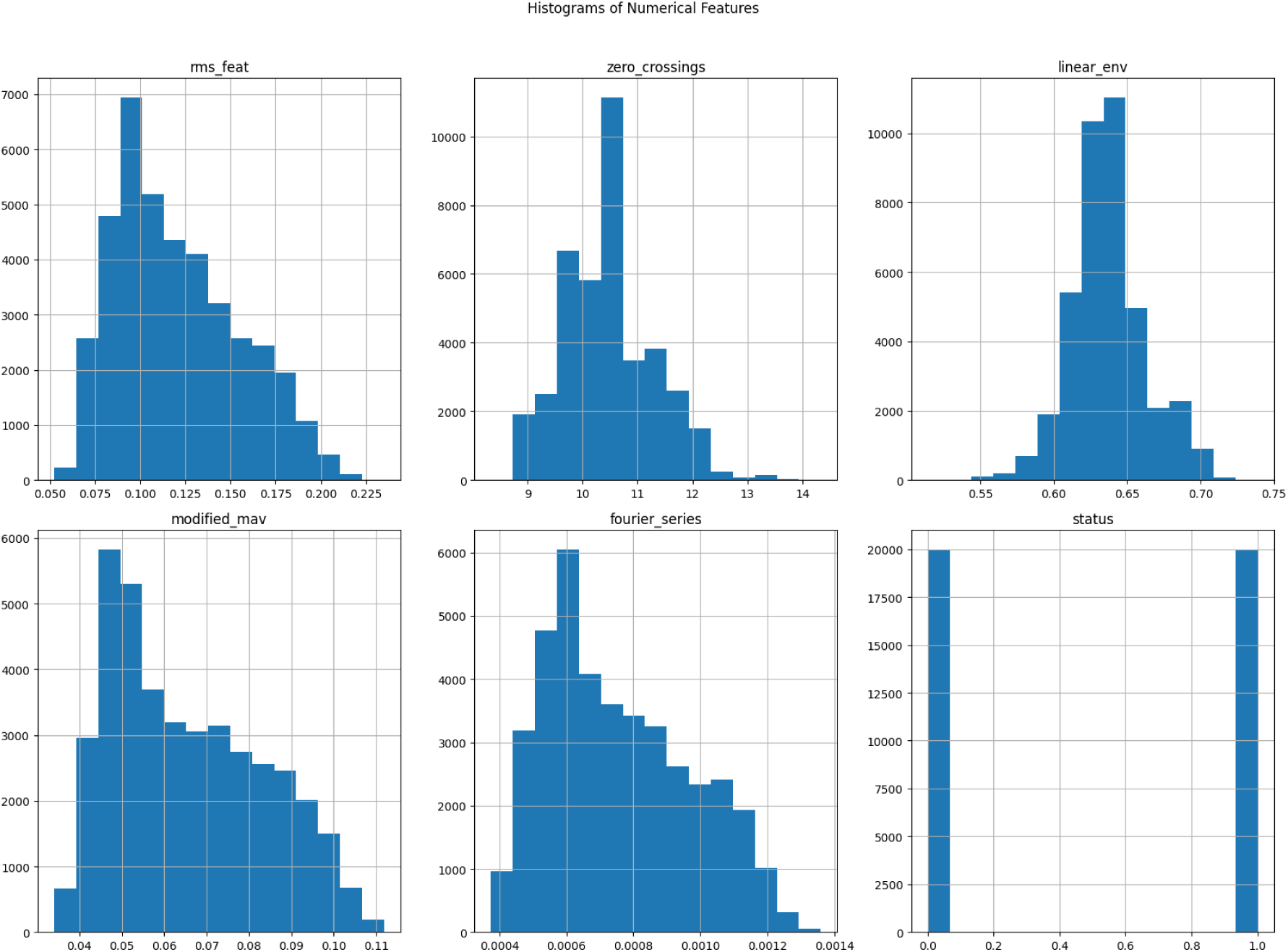
Histograms for Important Features generalize well on unseen data, making the model more robust.

### 2.7 Supervised Machine Learning

Since the dataset, as aforementioned, will be formed of various crucial features (16 features) supported with a label for each sample, supervised ML models were used. Studies indicate that the RF algorithm shows the highest accuracy among other tested algorithms when it comes to disease prediction and classifying medical big data [46] [47], and the SVM is most frequently used [46]. Therefore, for our study, we decided to use RF and SVM, in addition to Logistic Regression (LR), as a baseline model for the other methods and for the interpretability it brings. Moreover, RF, LR, and SVM do not assume that the input features need to be normally distributed, thus, in case of skewness, no effect will be encountered.

Data preprocessing included merging all the samples generated prior on a CSV file, checking for unintentional null values, and ensuring consistency of the features. One-third of the dataset was used for the testing data, and all models followed the same setup and training strategy. To reduce the risk of overfitting, *KFold Cross-Validation* is used where the data is split into “folds” (subsets). The model would be trained on the folds except for one used for the testing. This process is repeated until each fold is used as a test set once, thus, minimizing biases. We also use *GridSearchCV* to perform an exhaustive search over a grid of parameters defined earlier to find the best combination for the model eventually. It also integrates the KFold Cross-Validation while finding the best hyperparameter to

## 3 Results

To test the dataset, ML models were developed. After extracting these features, the models used to validate the dataset were RF, SVM, and LR. Eventually, after the models had been fitted, the scores, a classification report, and a confusion matrix were retrieved to evaluate each model’s performance. The scores in Table 2 show satisfactory results for each model’s best performance during the grid search process.

**Table 2:** Model Performance Comparison.

| Models Applied | RF | SVM | LR |
| --- | --- | --- | --- |
| Best Score (Grid Search) | 0.9817 | 0.8949 | 0.8324 |
| Accuracy | 0.9812 | 0.8997 | 0.8336 |
| Recall | 0.9831 | 0.8912 | 0.8353 |
| F1-Score | 0.9811 | 0.8984 | 0.8333 |

The RF algorithm resulted in the highest *best score (grid search)* of 98%. The SVM algorithm also achieved excellent results with a *best score* of 89%, while the LR algorithm had the lowest *best score* of 83%. Ultimately, the RF model has the best performance, while the LR performed the worst relative to the other two models’ performance. Nonetheless, the three models’ performance is considered quite satisfactory. It can also be further shown through the confusion matrices for each model (Table 3).

**Table 3:** Confusion matrices for the RF, SVM, and LR models.

| Random Forest |  |  | Support Vector Machine |  |  | Logistic Regression |  |  |
| --- | --- | --- | --- | --- | --- | --- | --- | --- |
|  | Predicted Positive | Predicted Negative |  | Predicted Positive | Predicted Negative |  | Predicted Positive | Predicted Negative |
| Actual Positive | 5900 | 125 | Actual Positive | 5471 | 554 | Actual Positive | 5012 | 1013 |
| Actual Negative | 101 | 5874 | Actual Negative | 650 | 5325 | Actual Negative | 984 | 4991 |

These results not only explain how successfully these algorithms performed but also validate the semi-synthetic dataset built and show the desired credibility. Despite the satisfactory results, a noted limitation is in the percentage of incorrect predictions, which is correlated to the edemas that are located on the lower bound (10%). Another limitation faced is the lack of publicly available datasets that show MRI scans of injured hamstring muscles. However, the developed approach made it possible to extend the dataset by extracting other boundary points from new MRI scans and thus generating additional EIT voltage files using EIDORS. The dataset and models can be found in the GitHub repository [48].

## 4 Discussion

In this study, a semi-synthetic dataset was developed to predict HSIs using EIT measurements. MRI scans were used to generate different cases of healthy and injured hamstrings, which allowed the developed dataset to cover different scenarios. Using EIDORS, a total of 120,000 EIT voltage files were created and constituted the dataset. Initial testing of the dataset on the raw EIT measurements using ML models resulted in poor performances and overfitting. Therefore, several features were extracted, which significantly improved the performance of the models, with the best-performing model being the RF with a 98% accuracy, followed by SVM and finally LR. These results validate the dataset and the ability of EIT to be used with the goal of distinguishing between injured and healthy hamstrings. This study lays the groundwork for future work aiming at predicting HSIs in real-time before their occurrence, especially for athletes. Achieving this goal would result in reducing injury rates which would potentially lead to an extended career period and a better performance.

## Data Availability

The original data presented in this paper is publicly available in the GitHub repository as previously cited in the paper and included here as a reference [48]. The repository includes the dataset and models needed to replicate the results of the paper.

https://github.com/reemshehayib/MUSCLE-Hamstring-Injury-Prediction-EIT

https://www.youtube.com/@dr.gayfirstlookmri

## Author Contributions

Conceptualization: N. Maalouf. MRI Scans Collection and Data Extraction: L. Youssef Baby. Dataset Creation: L. Youssef Baby and R. Shehayib. Machine Learning Models Development and Testing: R. Shehayib. Writing—original draft: L. Youssef and R. Shehayib. Writing—review and editing: N. Maalouf. Supervision: N. Maalouf. All authors have read and agreed to the published version of the manuscript.

